# Effects of Minimally Processed Red Meat Within a Plant-Forward Diet on Biomarkers of Physical and Cognitive Aging: A Randomized Controlled Crossover Feeding Trial

**DOI:** 10.1101/2025.08.11.25333443

**Authors:** Saba Vaezi, Bruna O. de Vargas, Jessica L. Freeling, Lee Weidauer, Moul Dey

**Affiliations:** School of Health and Human Sciences, South Dakota State University, Brookings, SD, USA; FreelingBio Research Consulting, LLC, Vermillion, SD, USA

**Author notes:** Correspondence: Moul Dey.

**Keywords:** minimally processed lean red meat, plant-forward diet, aging, insulin, cardiometabolic health, feeding study, cognitive decline

## Abstract

**Background:** Popular dietary patterns for cardiovascular and cognitive health such as the Mediterranean and MIND diets emphasize plant-based foods while limiting red meat intake. However, most research combines processed and unprocessed forms, limiting conclusions about unprocessed red meat.

**Objective:** To evaluate the effects of incorporating minimally processed lean red meat into a nutrient-dense, plant-forward, healthy dietary pattern on markers of aging-associated health decline.

**Methods:** This 18-week all-food-provided randomized controlled crossover feeding PRODMED2 trial tested an omnivorous red meat diet with 162g/d minimally processed pork (MPP) against a macronutrient- and energy-matched no-meat control diet with minimally processed lentils (MPL). Serum biomarkers relevant to metabolic-related cognitive and physical health were explored in 36 adults aged ≥65 years. Primary and secondary endpoints included five cardiovascular-related markers, 12 nutrition- and neurotransmitter-related measures, two metrics of body composition, and two muscular fitness outcomes. Data was analyzed using robust mixed effects models adjusted for covariates.

**Results:** Intervention diets were well tolerated, with high adherence. Improvements in cognitive related metabolic biomarkers were observed across both arms. Fasting insulin declined more after MPP (*p* < 0.001), with a corresponding increase in SPISE (*p* = 0.032), though between-group differences were not significant. HDL was higher post-MPP than post-MPL (*p* = 0.034). Body weight decreased in both arms (*p* < 0.05), with a smaller lean mass loss trend following MPP. Grip strength and chair-rise performance were maintained. Neuroactive metabolites and bioactive amino acid profiles shifted favorably in both arms.

**Conclusion:** These findings challenge the perception that red meat is broadly unsuitable for older adults. Including familiar foods like red meat, particularly in minimally processed form and within a healthy overall dietary pattern, may provide age-associated health benefits and improve adherence to plant-forward diets. These results have important implications for healthspan of older U.S. populations where red meat remains popular.

**Clinical Trial Registry:** Registered at www.clinicaltrials.gov as NCT05581953 and NCT06261775.

## Introduction

The U.S. population is rapidly aging, creating a demographic shift and increased healthcare burden due to age-related chronic diseases. Major health challenges associated with aging include cardiovascular diseases (CVD), sarcopenia-related loss of physical function, and cognitive decline. Nearly a quarter of community-dwelling Americans aged 65 years and older have poor health with an additional 1.3 million living in nursing homes, unable to maintain an independence [1–4]. Lifestyle approaches to improve the health span have the potential to delay the onset of aging-associated decline in health and independence and improve quality of life in older adults [5, 6].

Among age-related conditions, cognitive impairment, particularly dementia, poses a significant and growing concern. Dementia is a progressive neurodegenerative disorder characterized by cognitive decline and loss of functional independence, with Alzheimer’s disease (AD) being the most common form [7]. With limited options for early-diagnosis, prevention, and treatment, about 14 million older Americans are projected to have dementia by 2060 [8]. This underscores the urgent need for strategies that target modifiable risk factors.

Emerging evidence suggests that in addition to being a major contributor to CVD metabolic dysfunction may play a key role in the development of both cognitive and physical decline in older adults[9]. For example, metabolic syndrome, affecting ∼40% of older adults, has been shown to modulate epigenetic changes that increase the risk of type 2 diabetes and all-cause mortality [10]. Risk factors such as insulin resistance (IR) and obesity are strongly associated with age-related cognitive decline [11]. IR has received particular attention, as impaired brain glucose metabolism may precede clinical dementia symptoms by over a decade. Due to this pronounced link, AD has been referred to as “type 3 diabetes” [12]. In addition, since the anabolic role of insulin promotes muscle protein synthesis and glucose uptake, IR in aging is also closely linked to muscle loss, sarcopenia, and loss of physical strength [13].

Given these interconnections, lifestyle interventions that target metabolic health hold great promise for preserving both cognitive and physical function in older adults. Diet, physical activity and fitness, sleep, and social engagement are increasingly recognized as key determinants of dementia risk [14]. Among these, nutrition stands out for its influence on cognitive-related metabolic (cognometabolic) health [15, 16]. Diets rich in plant-based foods and healthy fats like the Mediterranean, DASH, and MIND diets, have been associated with reduced cognitive decline [17–20]. While these diets show promise, the use of both processed and unprocessed forms of red meat in research, along with mixed results across studies, points to the need for a deeper understanding of how red meat affects cognometabolic health. For example, a cohort study reported that replacing a daily serving of processed red meat with legumes reduced dementia risk by 19% [21], and that the consumption of meat is a risk factor for IR and type 2 diabetes across populations [22]. In contrast, a meta-analysis of intervention trials showed that unprocessed red meat intake did not affect weight gain or related metabolic conditions [23], and that red meat can even improve cognitive functionality under certain circumstances [24]. Randomized controlled trials examining the effects of 100% minimally processed lean red meat on cognometabolic wellbeing when consumed as part of healthy dietary patterns in older adults remain scarce.

As such, dietary patterns and active lifestyles that support insulin sensitivity may help reduce cognometabolic risk in aging populations as well as support physical function. Emerging evidence suggests that diet influences not only conventional nutritional pathways, but also neuroactive compounds involved in mood, cognition, and metabolic regulation [25]. Amino acid precursors such as tryptophan and tyrosine contribute to the synthesis of key neuromodulators, including serotonin, dopamine, GABA, melatonin, and kynurenines [25–28]. Other dietary components may also influence neuromodulator activities indirectly through effects on peripheral metabolism and gut–brain communications [29–32].

We conducted a randomized controlled crossover feeding trial in community-dwelling older adults. Each participant completed two 8-week dietary intervention phases, one testing a lean red meat diet featuring minimally processed pork (MPP) and the other a macronutrient- and energy-matched control diet based on plant-sourced primary proteins such as lentils, referred to as minimally processed lentil (MPL). We proposed that the addition of minimally processed lean red meat to a nutrient-dense, plant-forward dietary pattern aligned with the USDA macronutrient recommendations would enhance cognometabolic health and muscular fitness related to healthspan. Primary outcome markers associated with insulin sensitivity, iron status, and cognitive function were tested along with the chair stand test. A range of secondary markers related to cognometabolic health were also tested.

## Methods

This publication reports primary and related secondary outcomes from the Protein-Distinct Macronutrient-Equivalent Diets 2 (PRODMED2) randomized controlled feeding trial. The PRODMED2 trial was registered on October 12, 2022, at ClinicalTrials.gov (NCT05581953) and approved by the Institutional Review Board at South Dakota State University (IRB #2209010-EXP). Participant recruitment began shortly after trial registration. Unless otherwise specified, the work was carried out at South Dakota State University in Brookings, South Dakota, in accordance with the Declaration of Helsinki, and all participants provided written informed consent. Detailed information regarding the overall study design and recruitment protocols has been previously published [33].

### Study Design

This study was a randomized, controlled, two-arm crossover feeding trial in which all meals were provided to participants in a pre-portioned, ready-to-heat format to minimize intake variability. The protocol included in-person site visits and data collection at three time points: baseline and the end of each dietary intervention phase. Dine-in and food pickup were scheduled in addition to the data collection visits.

### Participants

Participants were randomized in a 1:1 ratio using a block design (block size of two for individuals or four for couples). Randomization was conducted by a study team member not involved in outcome assessments or later data analysis. Participants were assigned to either MPP or MPL interventions following randomization. Laboratory personnel responsible for processing biospecimens and collecting assay data were blinded to ID assignments.

The primary outcomes of the PRODMED2 trial were serum ferritin, homocysteine, insulin, and chair stand performance. Secondary outcomes included body weight, glucose, triglyceride, total cholesterol, high-density lipoprotein cholesterol (HDL), phosphatidylcholine, and grip strength normalized to body mass. Brain-derived neurotrophic factor (BDNF) and a panel of neuroactive and methylation-related metabolites were also assessed as exploratory outcomes to further support primary and secondary observations. These included phenylalanine, glycine, glutamic acid, tyrosine, kynurenine, γ-aminobutyric acid (GABA), tryptophan, and choline. The Single

Point Insulin Sensitivity Estimator Index (SPISE) was calculated based on collected data. All outcomes were selected for their relevance to cognometabolic indicators in older adults and their known responsiveness to short-term dietary interventions. No serious adverse events or potential harms were reported during either intervention phase.

A total of 36 community-dwelling older adults completed the PRODMED2 feeding trial. Initial eligibility was assessed via telephone screening, and individuals meeting preliminary criteria were invited for an on-site informational and screening visit. At this visit, trained staff conducted clinical measurements and structured interviews to assess health history, medication use, and lifestyle behaviors prior to potential enrollment and informed consent signing.

Eligible participants were adults aged 65 years or older, of any race, sex, education-level or marital status, and in generally good health as confirmed by a routine physical exam within the past year. Inclusion criteria included absence of medically diagnosed type 2 diabetes, body weight ≥110 pounds (∼50 kg), and reporting a habitual omnivorous dietary pattern without special dietary restrictions. Participants were required to consume only study-provided foods (with pork as the sole meat source), attend in-person visits, and abstain from alcohol, supplements, and non-study foods for the duration of the trial. Common prescription medication use was allowed, and participants were encouraged to maintain their habitual physical activity patterns throughout the study.

Exclusion criteria included substance use (tobacco and recreational drugs), medications affecting metabolism (e.g. steroids), recent dieting or weight loss, and diagnoses of major chronic conditions (e.g., cancer, diabetes, history of heart attack or stroke, hepatic or gastrointestinal disease). Participants were also excluded for diagnosed impaired kidney function, recent major surgery that would significantly impair their mobility, mental health concerns affecting compliance, or any reasons which would make them unable to attend study visits. Recruitment and screening were conducted between Fall 2022 through summer 2023 on a rolling basis.

### Dietary Intervention and Adherence

The intervention diets were designed to align with the 2020–2025 USDA Dietary Guidelines for Americans (DGA) for adults aged 51 and older for macronutrient distribution with a plant-forward emphasis. Every main meal (breakfast, lunch, and dinner) included plant foods with an average of 102 servings of plant foods (vegetables, fruits, grains) per week. A moderate amounts of dairy, eggs, and plant oils were included. Both diets were implemented in a fully controlled, all-food-provided format, using a 7-d rotating menu developed using Nutritionist Pro™ software (Axxya Systems). As primary proteins with >45%E of total protein, participants consumed either 5.7 oz or 162 g/d of minimally processed lean pork or an equivalent amount of protein from lentils for eight weeks, separated by a two-week washout period. To minimize confounding, alcohol, soy, beef, poultry, seafood, and artificial sweeteners were excluded. All meals and snacks were prepared by trained staff, portioned to the nearest gram, and provided either on-site or as take-home packages with clear heating instructions. Participants’ baseline diet reflected a typical omnivorous pattern, confirmed via 24-hour dietary recalls.

At the end of each dietary phase, participants completed a structured questionnaire assessing adherence, menu acceptability, and overall feasibility of the intervention. The survey included items on food acceptability, consumption of non-study food, self-reported compliance, convenience of meal delivery, likelihood of continuing a similar dietary pattern, and willingness to recommend the study to others. These responses captured key dimensions of participant satisfaction, adherence, and the potential for longer-term dietary behavior change.

### Sample Collection

Overnight fasted blood samples were collected at baseline and at the end of each dietary phase using standard venipuncture procedures. Blood was drawn into red-top tubes for serum and heparinized green-top tubes for plasma. Samples were anonymized, labeled, and immediately processed. Fresh whole blood was used for routine clinical chemistry analyses, including glucose and lipid panels. Serum was aliquoted and stored at –80°C for later batch analysis of circulating biomarkers, including biogenic amines and neuroactive metabolites. All procedures followed standardized protocols to ensure sample integrity and consistency across time points.

### Anthropometry, Body Composition, Muscle Function, and Blood Pressure

Anthropometric, body composition, and physical function measurements were conducted at baseline and at the end of each dietary intervention phase using standardized protocols. Standing height was measured to the nearest 0.5 cm using a wall-mounted stadiometer (Seca), and body weight was recorded to the nearest 0.1 kg using a calibrated digital scale (Seca) with participants in light clothing and no shoes. Body composition was assessed using dual-energy X-ray absorptiometry (DXA; Hologic Horizon), which provided estimates of fat-free mass (bone mass + lean mass), referred to as lean mass hereafter. Bone mass is unlikely to change within the short duration of the intervention; hence any observed change implies lean mass or muscle mass change.

Muscle strength and function were assessed using handgrip strength (measured as the highest value from three trials of the dominant hand using a handheld dynamometer) and the five-repetition chair rise test (time to complete five consecutive unassisted stands from a seated position), where longer completion times indicate poorer lower-body strength and functional performance.

Resting blood pressure was measured at baseline on the upper left arm using an automated sphygmomanometer (GE Carescape V100) after at least five minutes of seated rest. Two readings were taken one minute apart, and the average was used in the analysis.

### Biomarker Assessment

Cognometabolic biomarkers were evaluated using fasting serum samples. Total cholesterol, HDL, triglyceride, and glucose were measured via point-of-care testing using the Cholestech LDX System (Abbott Laboratories), in accordance with manufacturer guidelines. Serum concentrations of insulin, ferritin, and BDNF were quantified using magnetic bead–based multiplex immunoassays (MAGPIX, Luminex Corporation). These biomarkers are relevant to the health of older adults given their roles in CVD, IR, iron status, neuroinflammation, and neuroplasticity, all of which are increasingly recognized as interconnected factors influencing cognitive aging. Phosphatidylcholine plays a key role in membrane integrity, lipid metabolism, and neurotransmitter synthesis, making it relevant to both metabolic and cognitive health. Phosphatidylcholine and free choline levels were assessed using enzymatic colorimetric assays. Serum homocysteine, involved in one-carbon metabolism and associated with increased risk of CVD and cognitive decline, was measured using the Centaur CP system (Siemens). Batch analyses were performed at the Clinical and Laboratory Services for the Advancement of Science (CLASS) Laboratory at the University of Michigan.

Following our previously published protocols [34], biogenic amine profiling was conducted at the West Coast Metabolomics Center (University of California, Davis) using an untargeted HILIC-qTOF-MS platform. Serum samples were extracted, dried, and reconstituted before assaying biogenic amine metabolites including kynurenine, GABA, tyrosine, tryptophan, phenylalanine, glycine, and glutamic acid. Injections were onto a Waters Acquity BEH Amide column (1.7 μm, 2.1 × 150 mm) and separation was achieved using a gradient of LC-MS grade water and acetonitrile with ammonium formate and formic acid. Mass spectra were acquired in positive ion mode with high resolution, and metabolite identification was based on retention time and m/z values, verified against authentic standards. Quantification was performed using external standard curves and internal isotope-labeled controls. Data were processed with mzMine and Agilent MassHunter, and normalized using the SERRF algorithm. Final values were reported as calibrated relative intensities (RI) to reflect relative abundances.

### Cardiometabolic Risk Scores Calculation

Body mass index (BMI) was calculated as weight in kilograms divided by height in meters squared (kg/m²). Atherosclerotic cardiovascular disease (ASCVD) 10-year risk and estimated vascular age were both calculated using the Framingham Risk Score algorithm, which incorporates age, sex, total cholesterol, HDL, systolic blood pressure (SBP), treatment for hypertension, smoking status, and diabetes status. Vascular age was used as a surrogate marker for the biological burden of cardiometabolic risk [35]. SPISE was used to estimate insulin sensitivity and calculated as [36] SPISE = 600 x HDL^0.185^/TG^0.2^x BMI^1.338^. Indices were calculated using fasting values collected at baseline and post-intervention time points.

### Sample Size and Statistical Analysis

Most research on red meat fails to differentiate between processed and unprocessed forms. To address our hypothesis that incorporating minimally processed lean red meat within a healthy plant-forward diet would support cognometabolic health, we proposed markers of insulin sensitivity (insulin), cognitive decline (homocysteine), iron status (ferritin), as well as muscular fitness (chair stand performance). Sample size calculations for the PRODMED2 trial were based on the ability to detect a clinically meaningful difference in circulating homocysteine, which exhibited the greatest variability in effect size among the primary endpoints based on previously published data: homocysteine, insulin, ferritin, and chair stand performance. Based on this, a sample size of 12 participants per diet arm was estimated to provide 90% power to detect the expected difference at a two-sided alpha level of 0.05 (total probability of making a false positive type I error is 5%, split equally between both tails). To account for an anticipated overall attrition rate of approximately 25%, the target enrollment was increased to 15 participants per arm (*n* = 30 total). However, due to early observations, we intentionally exceeded the proposed enrollment target to ensure adequate statistical power at study completion. Against an anticipated attrition of 25% throughout the entire study, an unexpected 25% post-enrollment dropout before intervention start raised concerns about potential compliance (**Figure 1**). Additional dropouts or protocol nonadherence were anticipated, particularly given that the vegetarian feeding phase (control intervention) was culturally atypical for the Midwestern older adult cohort. Because of the 18-week intervention length and these uncertainties, recruitment continued until at least 12 participants in each arm had completed the study. By that point, 43 participants had already begun the intervention, and without a valid reason, we did not wish to remove them from the study.

**Figure 1.**
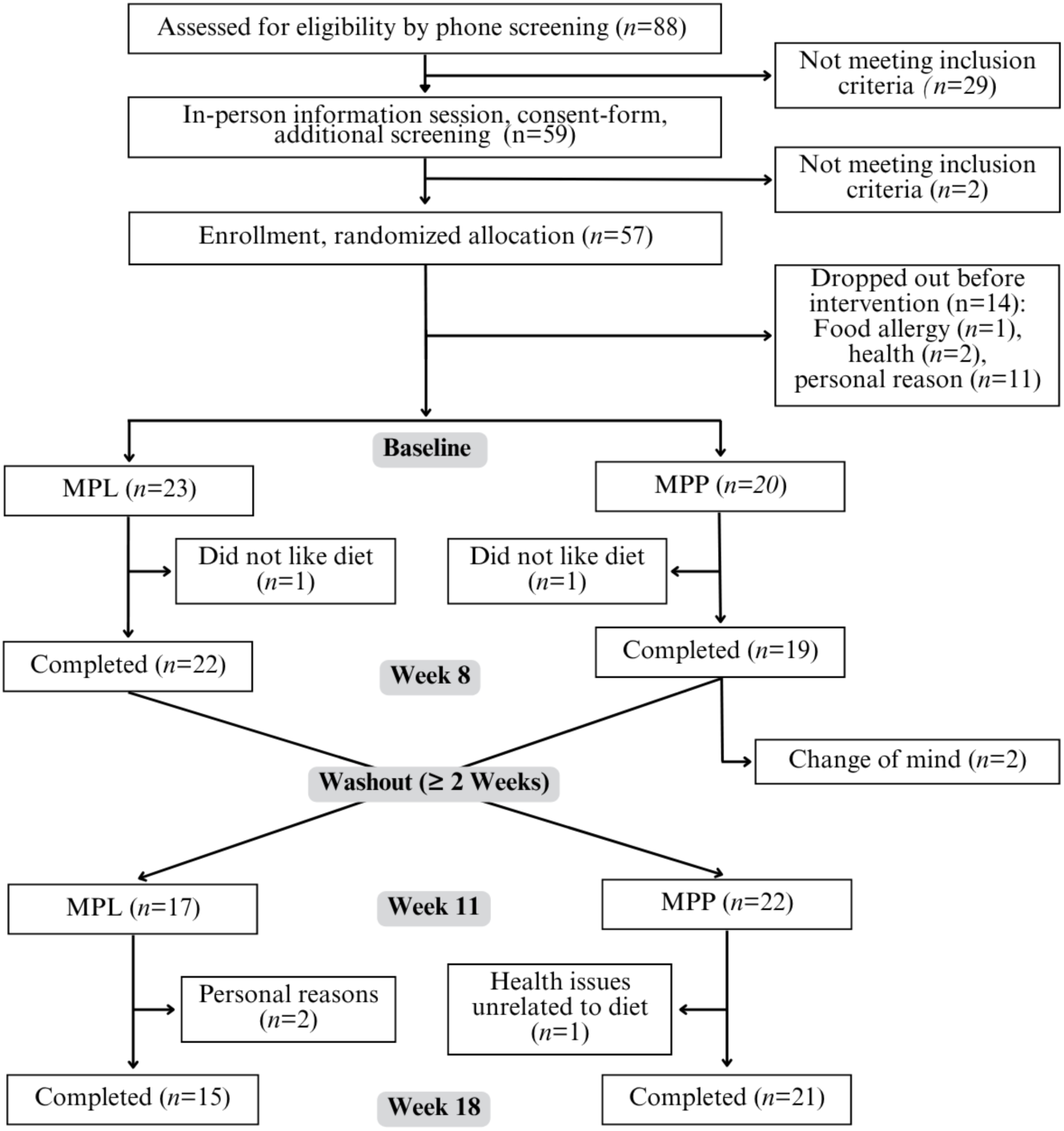
Flow chart and study design of the PRODMED 2 feeding trial. **Abbreviations**: MPP, minimally processed pork; MPL, minimally processed lentil; PRODMED 2, Protein-Distinct Macronutrient-Equivalent Diet 2.

All statistical analyses were performed using R version 4.3.2 (R Foundation for Statistical Computing) in RStudio. The normality of baseline variables was assessed using the Shapiro-Wilk test. Normally distributed variables were summarized as means ± standard deviations and compared using unpaired t-tests, while non-normally distributed variables were analyzed using Wilcoxon rank-sum tests. Descriptive and inferential analyses for primary and secondary outcomes were conducted using robust linear mixed-effects models (rLMMs) implemented with the robustlmm package; models included participant ID as a random effect and fixed effects for timepoint, age, and sex. Least square means and 95% confidence intervals (CIs) were estimated to evaluate biomarker differences across dietary phases. Pairwise comparisons were performed using the emmeans package with Tukey adjustment for multiple testing. For correlation analyses, Pearson’s or Spearman methods were used as appropriate. Visuals were generated using BioRender (BioRender.com), Adobe Illustrator 2025, and RStudio.

## Results

### Participant Characteristics

Of the 88 individuals screened, 57 met the eligibility criteria and were randomized to either MPP or MPL (**Figure 1**). Fourteen participants withdrew prior to baseline assessments mostly due to personal reasons (scheduling conflicts, obligations) or health-related exclusions. Among the 43 individuals who initiated the intervention (MPP: *n* = 20; MPL: *n* = 23), seven withdrew during the study period (MPP: *n* = 5; MPL: *n* = 2). A total of 36 participants who were initially assigned to MPP (*n* = 15) or MPL (*n* = 21) completed the full protocol and were included in the final analysis (**Figure 1**).

Baseline characteristics of participants are summarized in **Table 1**. The cohort was predominantly Caucasian older adults, with 72% female representation. Mean age was similar between sexes (females: 71.7 years; males: 71.8 years). Educational attainment was high, with over 70% of both males and females reporting a four-year college degree or higher. Nearly half of the participants were retired, and over half were married or living with a partner. Males were more likely to be married than females (80% vs. 50%).

**Table 1.**
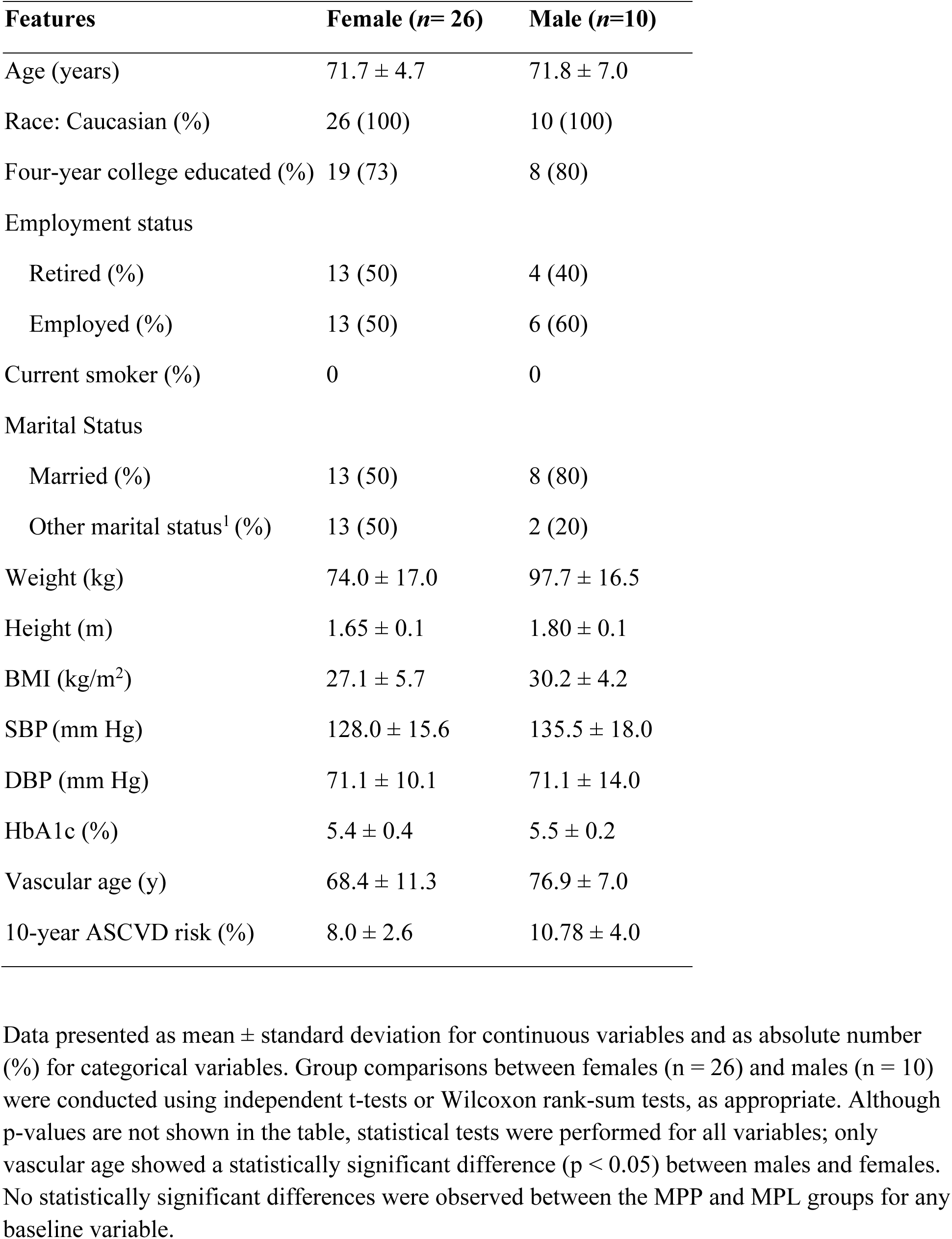
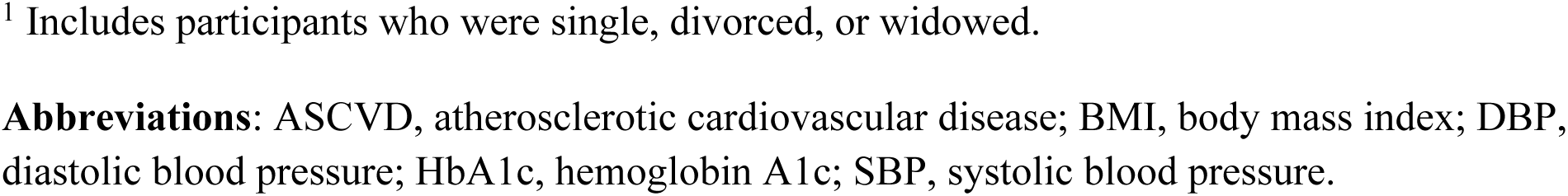
Baseline characteristics of the study participants by sex.

Participants demonstrated overall good health for their age, free of major chronic diseases but at risk for age-associated cognometabolic decline. Glycated hemoglobin values were within the normal range for both sexes (females: 5.4 ± 0.4%; males: 5.5 ± 0.2%). SBP was higher in males (135.5 ± 18.0 mm Hg) than females (128 ± 15.6 mm Hg), though sex difference was not statistically significant. Diastolic blood pressure (DBP) was identical between sexes (71.1 mm Hg). BMI was higher in males (30.2 ± 4.2 kg/m²) than in females (27.1 ± 5.7 kg/m²), though this difference did not reach statistical significance. Based on mean BMI, females were classified as overweight and males as obese. Despite identical chronological age, vascular age was significantly higher in males (76.9 ± 7 years) than in females (68.4 ± 11.3 years; *p* males vs females = 0.019). The estimated 10-year ASCVD risk score remained below 11% in both sexes.

### Nutritional Characteristics and Compliance of Provided Diets

All meals were fully provided, ensuring consistent dietary intake across participants. Energy intakes were similar between intervention phases, ranging from 1982.6 kcal at baseline to 2068.3 kcal in MPP and 2021.8 kcal in MPL (**Table 2**). Protein content comprised approximately 17– 18% of total energy. Compared to baseline, both diets provided similar energy, lower fats and saturated fats, and increased carbohydrates typical of a plant-forward dietary pattern. The provided diets met the Recommended Dietary Allowance (RDA) for B vitamins, including B6, B9 (folate), and B12. Self-reported adherence and satisfaction were high (**Figure 2**). Over 75% of participants reported high compliance. The convenience of pre-portioned meals was appreciated (MPP: 83%; MPL: 86%) and more than 50% of participants in both diets reported no consumption of non-study food. A majority indicated interest in continuing a DGA-aligned diet post-intervention. More MPP participants (78%) reported they would recommend the study, compared to 72% in the MPL group. Greater cultural familiarity with an omnivorous lifestyle in the Midwest and among this older adult population may have contributed to this survey outcome.

**Figure 2.**
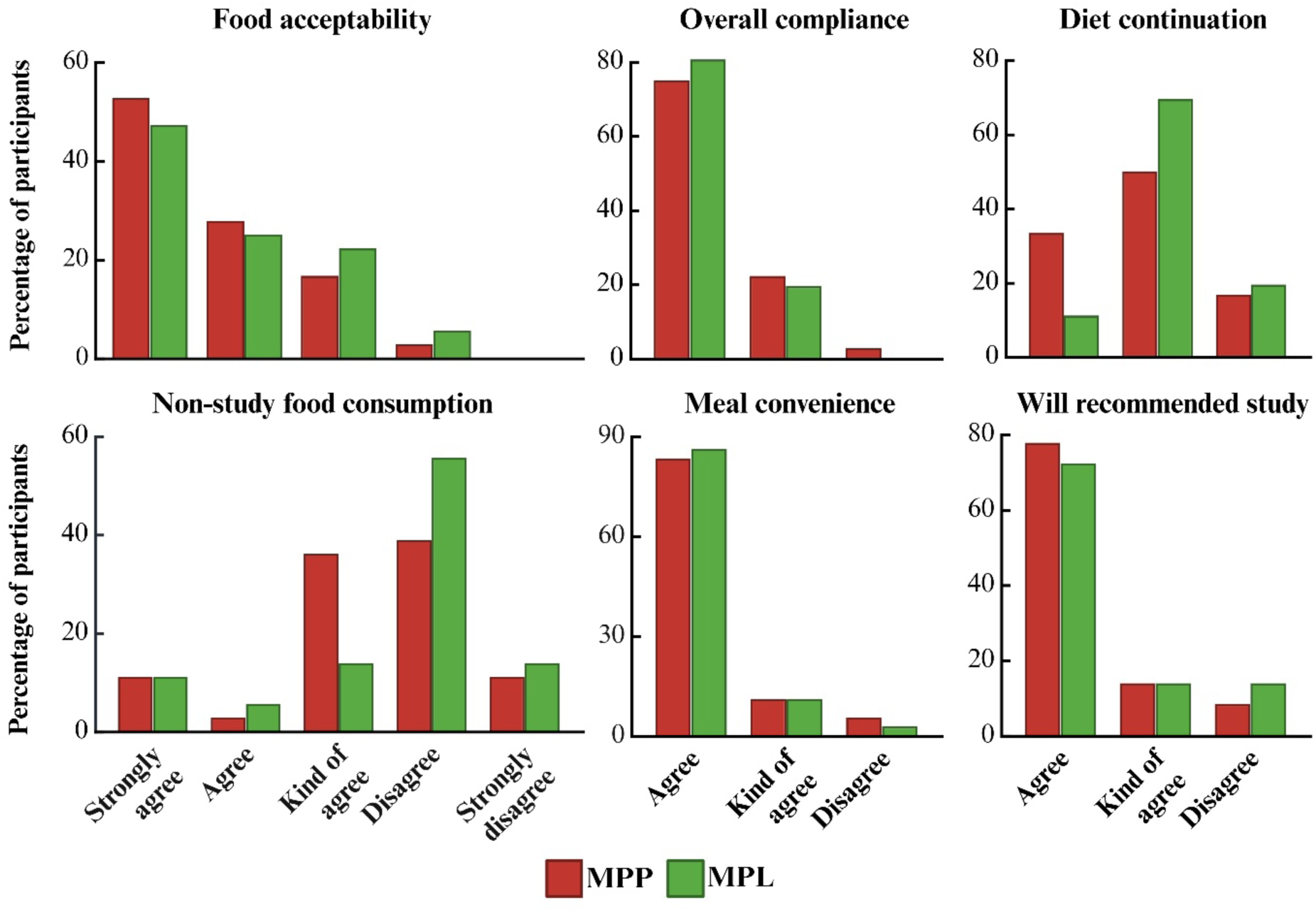
Participant-reported satisfaction and compliance with both diet phases. Responses are shown as the percentage of participants in each response category. **Abbreviations**: MPL, minimally processed lentil; MPP, minimally processed pork.

**Table 2.** Nutrient composition of the diets at baseline, MPP, and MPL phases.

| **Components** | **Baseline** | **MPP** | **MPL** |
| --- | --- | --- | --- |
| Energy (kcal/d) | 1982.6 | 2068.3 | 2021.8 |
| Protein (%E) | 17.2 | 18.0 | 17.2 |
| Carbohydrate (%E) | 43.2 | 55.7 | 54.2 |
| Fat (%E) | 41.6 | 29.4 | 30.7 |
| Saturated Fat (%E) | 12.9 | 6.8 | 8.7 |
| PUFA (g/d) | 15.2 | 21.2 | 17.7 |
| MUFA (g/d) | 33.5 | 12.4 | 13.8 |
| Sodium (mg/d) | 2548.5 | 2117.6 | 2194.9 |
| Vitamin B6 (mg) | 1.6 | 3.34 | 1.9 |
| Folate (mcg DFE) | 335.5 | 489.4 | 456.5 |
| Vitamin B12 (mcg) | 10.3 | 2.4 | 2.6 |
| Choline (mg) | 316.3 | 361.6 | 260.3 |

Baseline data (n = 35) are presented as mean daily energy and macronutrient intake, along with the provided values during the MPP and MPL diet phases. Macronutrient percentage distribution (%E) was calculated based on total energy provided.
**Abbreviations:** MPL, minimally processed lentil; MPP, minimally processed lean pork; %E, percentage of total energy; MUFA, monounsaturated fatty acids; PUFA, polyunsaturated fatty acids.

### CVD-related Biomarker Changes

Metabolic health is a critical component of healthy aging and is increasingly linked to cognitive health outcomes through shared mechanisms like insulin sensitivity, (e.g. cognometabolic health). To determine whether the intervention diets produced systemic effects on metabolic regulation, we examined circulating markers of glucose control, insulin sensitivity, and lipid profiles. Both diets were associated with favorable changes to markers of cognometabolic health relative to baseline, though the magnitude of response varied across markers (**Table 3**). Glucose declined significantly in MPL (−5.1 mg/dL, *p* = 0.002), with a similar but nonsignificant reduction in MPP (−3.4 mg/dL, *p* = 0.087). Fasting insulin concentrations decreased in both diet phases (both, *p* < 0.001), suggesting improved insulin sensitivity across both plant-forward diets irrespective of primary protein source. No significant between-group differences were observed for either glucose or insulin (*p* > 0.05). Insulin sensitivity was further assessed using the SPISE index, a non–insulin-based surrogate that incorporates lipid parameters. SPISE increased significantly following the MPP phase (*p =* 0.032), but not MPL (*p* = 0.269), suggesting a possible diet-specific enhancement in metabolic efficiency.

**Table 3.**
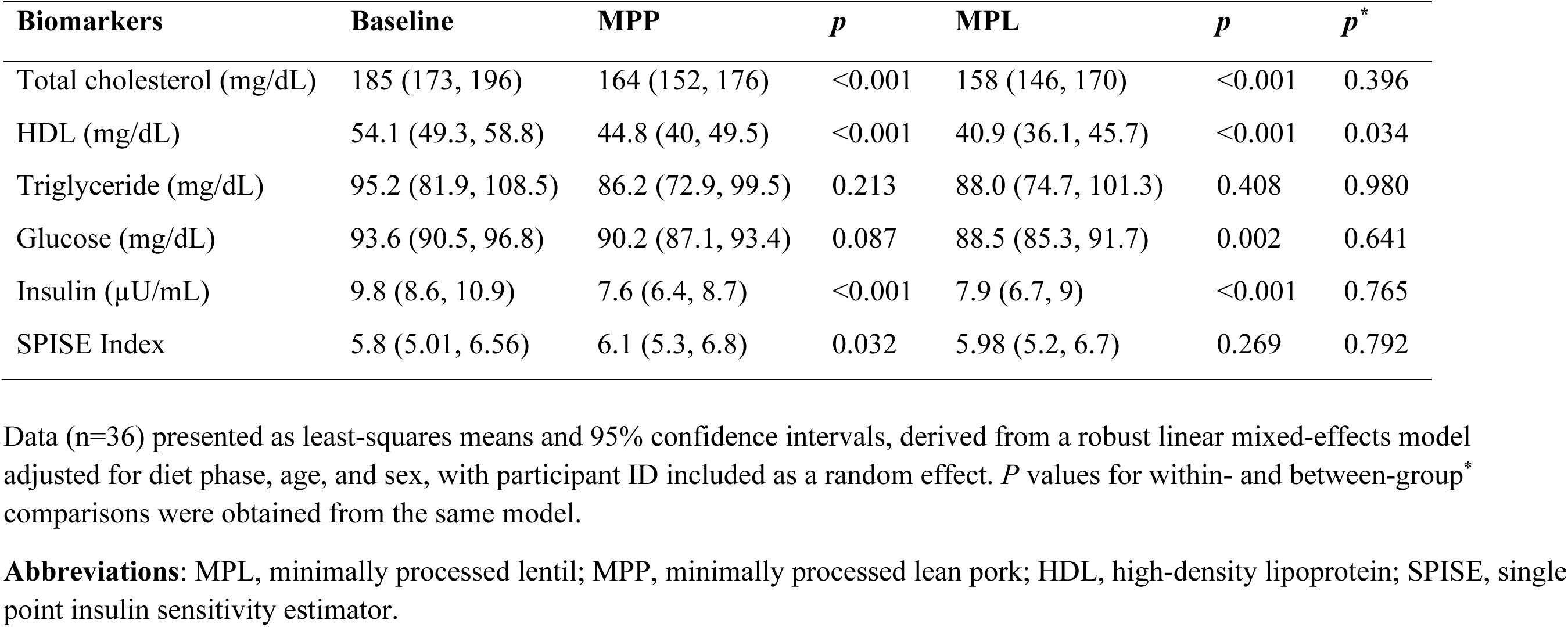
Cardiometabolic biomarker responses at baseline and during diet phases.

| **Biomarkers** | **Baseline** | **MPP** | ***p*** | **MPL** | ***p*** | ***p*<sup>\*</sup>** |
| --- | --- | --- | --- | --- | --- | --- |
| Total cholesterol (mg/dL) | 185 (173, 196) | 164 (152, 176) | <0.001 | 158 (146, 170) | <0.001 | 0.396 |
| HDL (mg/dL) | 54.1 (49.3, 58.8) | 44.8 (40, 49.5) | <0.001 | 40.9 (36.1, 45.7) | <0.001 | 0.034 |
| Triglyceride (mg/dL) | 95.2 (81.9, 108.5) | 86.2 (72.9, 99.5) | 0.213 | 88.0 (74.7, 101.3) | 0.408 | 0.980 |
| Glucose (mg/dL) | 93.6 (90.5, 96.8) | 90.2 (87.1, 93.4) | 0.087 | 88.5 (85.3, 91.7) | 0.002 | 0.641 |
| Insulin (μU/mL) | 9.8 (8.6, 10.9) | 7.6 (6.4, 8.7) | <0.001 | 7.9 (6.7, 9) | <0.001 | 0.765 |
| SPISE Index | 5.8 (5.01, 6.56) | 6.1 (5.3, 6.8) | 0.032 | 5.98 (5.2, 6.7) | 0.269 | 0.792 |

Data (n=36) presented as least-squares means and 95% confidence intervals, derived from a robust linear mixed-effects model adjusted for diet phase, age, and sex, with participant ID included as a random effect. *P* values for within- and between-group\* comparisons were obtained from the same model.
**Abbreviations:** MPL, minimally processed lentil; MPP, minimally processed lean pork; HDL, high-density lipoprotein; SPISE, single point insulin sensitivity estimator.

Lipid profiles improved under both dietary conditions. Total cholesterol decreased significantly in both MPP (−21 mg/dL) and MPL (−27 mg/dL) phases (*p* < 0.001), with no significant difference between groups (*p* > 0.05). HDL declined across both diets (*p* < 0.001); however, the reduction was smaller in the MPP group (*p* = 0.034 vs. MPL). Triglyceride levels reduced by a small margin in both groups, but the reductions were statistically non-significant. These results demonstrate that both primary protein sources improved multiple metrics of metabolic functions known to be associated with cognitive wellbeing in aging.

### Neurotransmitter and Methylation Pathway-Related Biomarkers

To explore diet-related effects on age-associated cognitive function, circulating biomarkers of neurotransmitter biosynthesis, methylation activity, and neurotrophic support were measured, reflecting neurochemical signaling and one-carbon metabolism (**Table 4**). BDNF, a key regulator of synaptic plasticity and neuronal health, increased modestly following MPL (*p* = 0.056) and showed no change in MPP suggesting no adverse effect of pork on circulating BDNF levels.

**Table 4.**
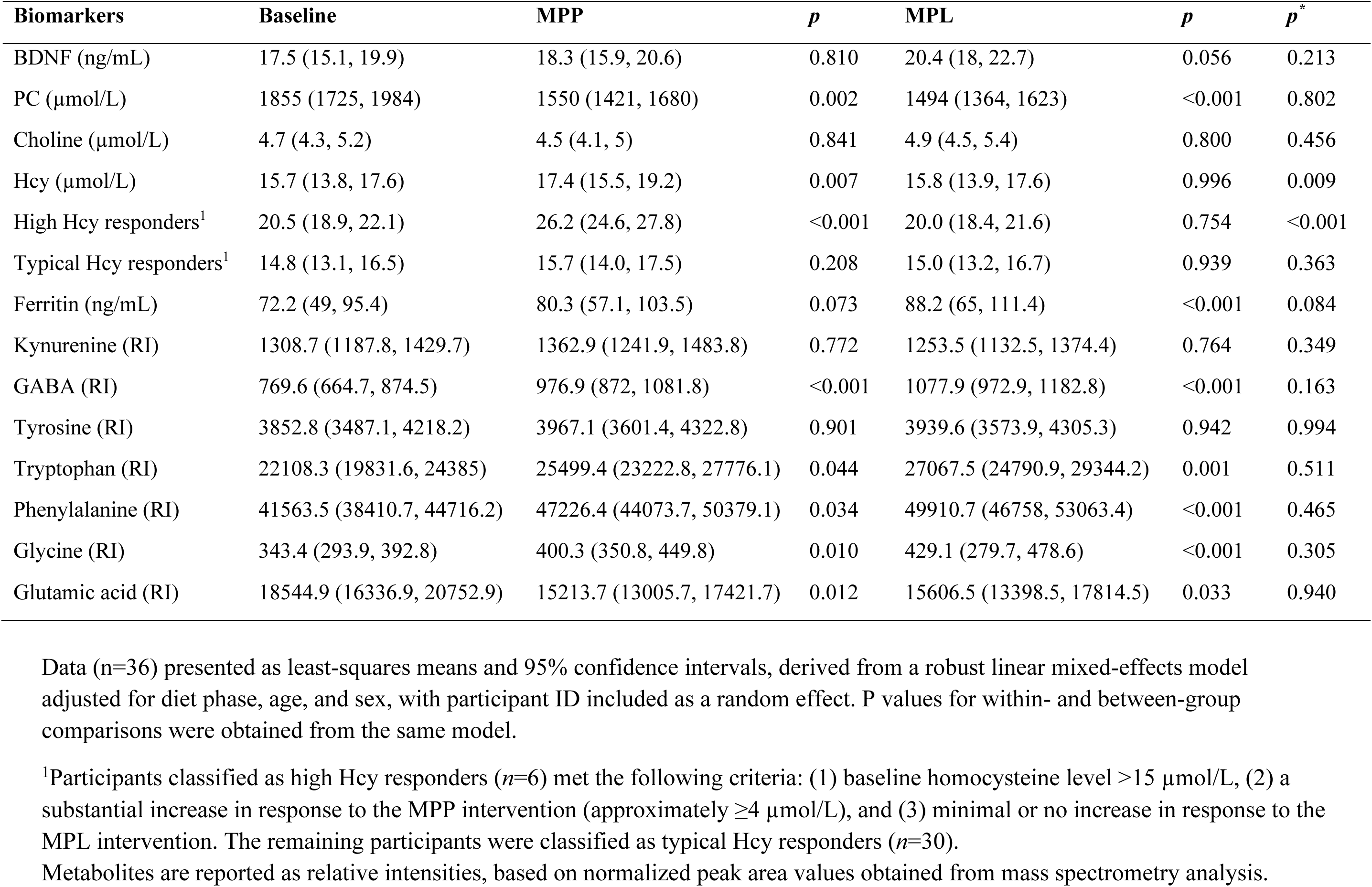
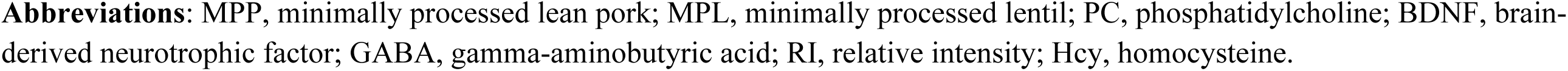
Effects of interventions on blood biomarkers of nutrition and neurotransmitter-related compounds.

Serum choline, a precursor to acetylcholine and phosphatidylcholine, remained stable across both diet phases, despite lower dietary supply indicating effective physiological regulation of this essential nutrient (**Table 2, 4**). In contrast, phosphatidylcholine levels declined significantly from baseline in both MPP (*p =* 0.002) and MPL (*p* <0.001), with no significant difference between groups (*p* = 0.802). This may reflect shifts in membrane lipid turnover or utilization in response to dietary composition.

Homocysteine, a key intermediate in one-carbon metabolism, increased modestly following the MPP phase (+1.7 µmol/L, *p* = 0.007), whereas it remained unchanged during the MPL phase (*p* = 0.996), resulting in a between-group difference (*p =* 0.009) (**Table 4**). Although homocysteine levels tend to be higher in older adults compared to younger individuals, the commonly accepted threshold for elevated homocysteine remains ∼15 µmol/L across all age groups [37]. Using this threshold, half of the participants (*n* = 18) exhibited >15 µmol/L baseline homocysteine levels. Among these, six participants (16.6%) showed a markedly different response to the MPP intervention compared to MPL: their homocysteine levels remained stable after MPL (*p* = 0.754) but rose substantially after MPP (+5.7 µmol/L from an already high baseline of 20.5 µmol/L, baseline to post-MPP, *p* = <0.001; post-MPL vs. post-MPP, *p* = <0.001). For the remaining participants (*n* = 30), the mean baseline homocysteine level was 14.8 µmol/L and remained stable after both MPP and MPL diets (baseline to post-MPL, *p* = 0.939; baseline to post-MPP, *p* = 0.208; post-MPL vs post-MPP, *p* = 0.363). Homocysteine levels in the six hyper-responders to MPP remained substantially higher than in the other 30 participants at all time points (all p ≤ 0.004), regardless of diet. Serum vitamin B12 levels were 22–33% lower in the hyper-responders than in the rest of the cohort at all time points, reaching statistical significance only after MPP (*p* = 0.041), when homocysteine levels were highest. However, all participants consistently maintained serum vitamin B12 concentrations above the clinical reference range The published reference range of serum vitamin B12 for adults is generally 160–950 pg/mL [38].

Ferritin was examined for its role in brain iron regulation, oxidative stress buffering, and neuroprotection. Ferritin levels rose in both groups (+8.1 ng/mL post-MPP vs +16 ng/mL post-MPL) reaching statistical significance in MPL (*p* < 0.001) and trending toward significance in MPP (*p* = 0.073), with no significant difference between diets (*p* = 0.084) (**Table 4**).

Several neuroactive metabolites were also responsive to dietary intervention. Both groups exhibited significant increases in GABA (MPP: *p* <0.001; MPL: *p* < 0.001), alongside corresponding reductions in glutamic acid, its excitatory precursor (MPP: *p* = 0.012; MPL: *p* = 0.033), with no between-group differences (*p =* 0.940). These changes may reflect a shift toward greater inhibitory neurotransmitter activity, potentially supporting neuronal homeostasis.

Within the serotonin pathway, tryptophan levels increased significantly in both groups (MPP: *p* = 0.044; MPL: *p =* 0.001), while kynurenine remained unchanged. Phenylalanine and tyrosine, both precursors in dopamine biosynthesis, increased, with phenylalanine reaching significance in both groups (MPP: *p* = 0.034; MPL: *p* <0.001). Glycine, a co-agonist at N-Methyl-D-aspartic acid (NMDA) receptors that mediate excitatory neurotransmission, increased significantly in both phases (MPP: *p* = 0.010; MPL: *p* < 0.001). Taken together, these findings suggest that both diets modulated circulating metabolites involved in neurotransmission and one-carbon metabolism, representing early biochemical adaptations with potential relevance to neurochemical balance, brain health, and cognitive resilience during aging (**Table 4**).

### Changes in Body Composition and Physical Function

Body composition and physical function are integral to age-related well-being and independence and are closely interconnected with cognometabolic functionality through shared pathways involving insulin signaling and nutrient metabolism. We assessed the effects of the dietary interventions on body weight, tissue mass distribution, and performance-based outcomes in the absence of a physical activity intervention component. As shown in **Figure 3A**, total body weight significantly decreased following both MPP and MPL diet phases compared to baseline (MPP: –4.6 kg; MPL: – 5.1kg; both *p* < 0.001), with no significant difference between diets (*p* > 0.05). With weight loss, lean mass loss may be unavoidable without a substantial strength training intervention, which was not included in this study. However, lean mass loss was lower post-MPP (–1.2 kg, *p* < 0.001) than post-MPL (–1.6 kg, *p* < 0.001), although not statistically significant between the diets (*p* = 0.249) (**Figure 3B**), suggesting a potentially modest benefit of minimally processed pork intake in attenuating age-related muscle loss.

**Figure 3.**
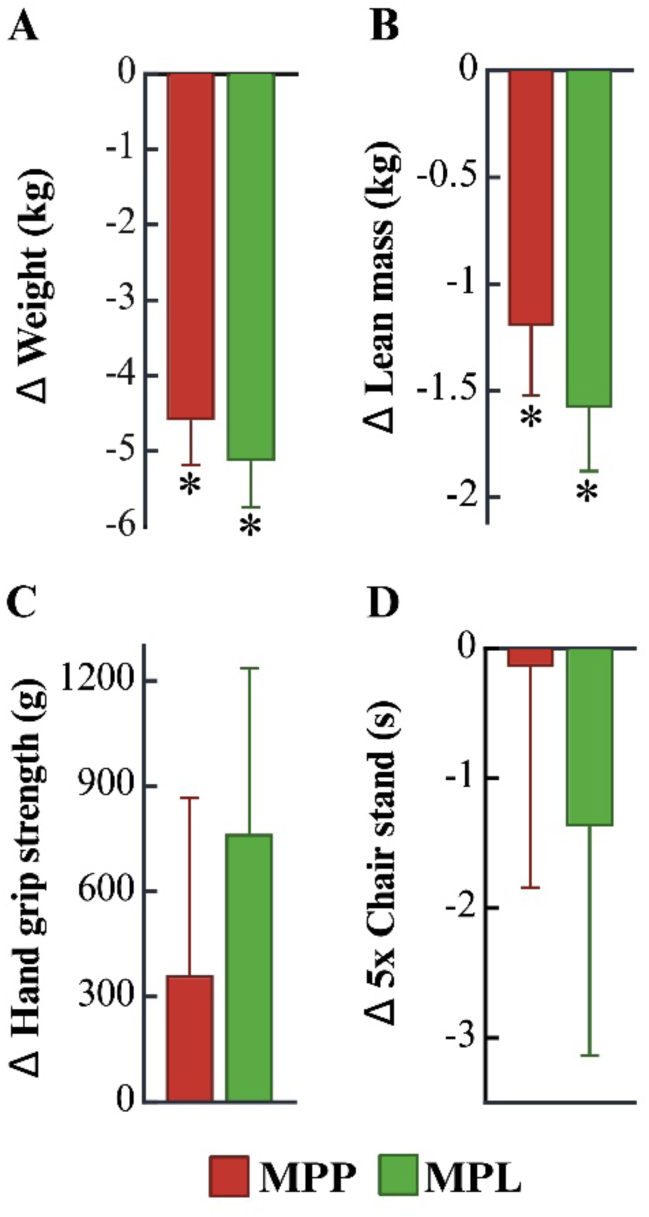
Body composition and physical function outcomes following each diet phase. Changes in body weight (A), lean mass (B), handgrip strength (C), and chair stand performance (D) across baseline and intervention diets. Change data post-intervention are presented as mean ± SEM with reference to baseline for participants with available measurements (n = 36 for all outcomes, except end-of-study body weight, n = 35; chair stand performance, n = 32 due to four participants unable to perform the test). Asterisks (*) indicate significant within-group changes from baseline (p < 0.05), based on robust linear mixed models. **Abbreviations**: MPP, minimally processed lean pork; MPL, minimally processed lentil.

Functional outcomes are presented in **Figure 3C** and **D**. Handgrip strength remained stable across all time points, with minimal improvements but no decline from baseline (*p* > 0.05, between groups). Lower-body function, assessed via the five-repetition chair stand test, also remained stable following both interventions with no signs of decline (all, *p* > 0.05).

## Discussion

As the number of older adults in the population continues to increase, metabolic-associated neurodegenerative diseases are becoming increasingly prevalent. This randomized controlled feeding intervention evaluated the effects of incorporating minimally processed lean pork daily within a plant-forward nutrient-dense healthy dietary pattern on clinical and biomarker-based outcomes related to age related cognometabolic functions. In this cohort of older adults with generally healthy baseline profiles for their age, the differences between the presence or absence of red meat in their diet were small. Both dietary interventions led to comparable improvements in insulin sensitivity and other cognometabolic features including circulating neuroactive amino acids, whereas functional fitness-related outcomes were preserved without any strength training intervention. Notably, the absence of adverse health effects from daily minimally processed red meat intake, challenges the common perception that red meat should be broadly restricted.

Both diets improved fasting insulin and total cholesterol which are independent risk factors for CVD and dementia-related illnesses [39]. The MPP diet notably enhanced the SPISE index, indicating improved insulin sensitivity. While the total cholesterol decreased, a modest decline in HDL cholesterol was also observed in both groups; however, the reduction was smaller in the MPP group. Body weight decreased with both plant-forward diets, and there was a trend toward less lean mass loss following the pork-based intervention. This pattern is consistent with an RCT finding that pairing resistance training with a lean red meat diet boosted muscle and strength in older women, supporting healthy aging [40]. While observational data often links red meat with adverse metabolic outcomes [41–43], a meta-analyses of 21 randomized controlled trials found no consistent negative effects of red meat intake on glycemic or insulinemic risk factors for type 2 diabetes [44]. Similarly, pooled analyses report no significant differences in CVD risk markers when red meat, especially the unprocessed form [23], was compared with a variety of other dietary proteins. In some cases, red meat consumption has even been associated with improvements in lipid parameters when compared with carbohydrates, mixed animal proteins, or habitual diets [45–47], all of which are in alignment with the observations from this RCT.

Beyond cognometabolic risk factors sharing pathways between AD and CVD, neurotransmitter dysfunction is a key feature of AD that contributes to both cognitive behavioral symptoms related to age associated mental health decline. For example, reduced GABA serum levels have been linked to psychological disturbances [48]. In our study, GABA increased, and glutamic acid decreased with both interventions, suggesting a shift toward improved inhibitory balance. The serotonergic system, also disrupted in AD, plays a critical role in memory and mood [49]. Tryptophan, an essential amino acid precursor for serotonin, increased with both diets. But tryptophan’s metabolite, kynurenine remained unchanged, indicating preserved serotonergic support. For dopamine-related pathways, phenylalanine increased across both diets, while tyrosine remained stable aligning with existing research [50] as potentially beneficial. Additionally, glycine, a co-agonist at NMDA receptors, rose modestly and may contribute to enhanced synaptic plasticity and cognitive function [51]. Although not reflective of central levels, these shifts in circulating neuroactive amino acids suggest that a plant-forward dietary pattern may influence peripheral markers associated with brain health. The inclusion of red meat did not diminish these potential benefits in this cohort of older adults. Importantly, these neuroactive biomarker shifts occurred in parallel with favorable cognometabolic and cardiovascular outcomes, underscoring the need to evaluate red meat within the context of the overall dietary pattern rather than in isolation.

Some studies emphasize the relevance of the dietary matrix and the nature of the comparison diet when assessing red meat’s health effects [43]. Our findings within the context of a plant-forward healthy dietary pattern, support the idea that lean, minimally processed red meat can be consumed regularly without adverse cognometabolic consequences. This has important public health implications; particularly for older adults in rural Midwestern communities where red meat is not only a cultural staple but may also serve as a familiar and acceptable component that facilitates the adoption and long-term adherence to healthier plant-forward dietary patterns.

While several outcomes showed clear benefits of red meat intake from baseline to post diet, other observations were more nuanced, pointing to potential areas of follow-up research. A modest increase in circulating homocysteine was observed in the MPP group but not in the MPL group, when all 36 participants were considered. Homocysteine levels typically increase with age and show an inverse correlation with B12 bioavailability. Intake of vitamin B12 which plays a critical role in homocysteine remethylation and clearance [31], remained above-RDA level during the interventions but was generally lower than baseline intake levels. A study reported that a 9 to 20 μmol/l blood homocysteine range may be indicative of lowest mortality rate in elderly patients [52]. The overall increase in homocysteine following MPP was slightly above the reported 16.5 μmol/L physiological range for this age group and well below the 20 μmol/L mark [53]. However, since homocysteine is recognized as a potential marker of cognitive decline [54], we further explored individual variability. Notably, six participants with elevated baseline homocysteine levels exhibited a distinct increase in response to red meat intake, unlike the remaining 30 participants whose levels remained stable. This suggests a possible subpopulation effect, which may reflect individual variability in one-carbon metabolism or micronutrient status and warrant further investigation. Studies have shown that some older adults may have a reduced ability to absorb food-bound B12 due to which they exhibit hyperhomocysteinemia in general or in response to higher methionine containing foods such as red meat [55–58]. The six hyper-responders consistently had much lower average concentrations of serum B12 levels than the remaining 30 participants across all time points.

While the physiological risks and benefits of red meat have been widely studied and debated in general, the specific effects of lean, unprocessed red meat consumed within a healthy dietary matrix on metabolic and neuroactive risk factors remain poorly understood. Red meat has been linked to elevated homocysteine, although findings are mixed, with one multicenter study reporting that refined cereal consumption was more strongly associated with circulating homocysteine levels than red meat intake [59–61]. Based on our homocysteine data, we speculate that a precision nutrition approach may help address many inconsistencies. Large scale studies are needed to clarify whether red meat, when consumed within an overall healthy dietary pattern, supports chronic disease management for most individuals, but may be less suitable for a subset of older adults with distinct metabolic profiles. This view is supported by animal studies showing that altering gut microbiota composition eliminated a diet-induced high-homocysteine phenotype [62].

Additionally, phosphatidylcholine, a key dietary and membrane-bound source of choline and a precursor for acetylcholine synthesis [63], declined following both dietary interventions. Although free choline levels were maintained, likely due to tight homeostatic regulation, choline intake remained below recommended levels across all time points. The parallel decline in phosphatidylcholine across both groups may reflect a compensatory mechanism [64], rather than a deficiency *per se*. However, this finding raises the possibility of a marginal choline insufficiency, which could be relevant in the context of plant-forward diets for older adults. While not conclusive, this aligns with existing evidence that choline intake is often suboptimal in the general population [65], and that supplementation may be warranted.

Our study design and implementation protocol had several strengths and general limitations that were separately published [33]. Overall, a feeding study is logistically challenging but offers stronger evidence for causal treatment effects compared to observational studies. However, feeding trials evaluating the effects of minimally processed red meat on cognometabolic risk factors are especially scarce. A specific limitation of this work relates to the relatively shorter duration of the intervention, due to which functional cognitive testing was not included. We believed that functional cognitive changes would be less likely to emerge within the relatively short 8-week intervention period. A second limitation is that the study population consisted exclusively of Caucasian adults, reflecting the demographic makeup of the rural Midwestern region where the research was conducted. As a result, caution is warranted when applying these findings to more diverse populations, including those of different racial backgrounds, age groups, body compositions, or with varying health conditions.

## Conclusion

Findings from this randomized controlled crossover feeding trial in rural Midwestern older adults suggest that minimally processed lean red meat such as pork, when incorporated into a plant-forward diet, can support cognometabolic health, help preserve muscle function, and may influence peripheral neurochemical markers. Supporting healthy aging may be better achieved through nutrient-dense, and balanced diets that draw on high-quality foods from both plant and animal sources. This dietary pattern reflects locally relevant food practices in a Midwestern population where red meats and healthy plant-based foods are widely available and commonly consumed. Further research is needed to assess the long-term clinical relevance of these findings.

Author contributions

The authors’ responsibilities were as follows: MD conceived the project, designed the research, and provided resources and study oversight; MD, BOdV, SV, and LW conducted the research and collected data; SV analyzed and visualized the data; SV, BOdV, JLF, and MD contributed to data annotation and interpretation; MD and SV wrote the manuscript and hold primary responsibility for the final content. All authors read, helped edit, and approved the final manuscript.

## Acknowledgements

The authors gratefully acknowledge following data collection support in an investigator-blinded manner: from the West Coast Metabolomics Center at the University of California, Davis, for conducting metabolomics assays and providing standard curve–based untargeted quantitative data (NIH U2C ES030158), and from the Central Ligand Assay Satellite Services Laboratory at the University of Michigan School of Public Health for certain biomarker panels. The authors also extend their sincere appreciation to the clinical research staff at South Dakota State University for their essential support in day-to-day logistics while implementing the feeding trial as well as express heartfelt thanks to all study participants for their time and commitment to the research.

## Data availability

Data is available upon reasonable request by contacting the corresponding author.

## Declaration of Generative AI and AI-assisted technologies in the writing process

During the preparation of this work, the authors used ChatGPT to assist with sentence structure and grammar. All content was subsequently reviewed and edited by the authors, who take full responsibility for the final version of the manuscript.

## Abbreviations

AD: Alzheimer’s disease
ASCVD: atherosclerotic cardiovascular disease
BDNF: brain-derived neurotrophic factor
BMI: body mass index
cognometabolic: cognitive-related metabolic health
DGA: Dietary Guidelines for Americans
DXA: dual-energy X-ray absorptiometry
GABA: gamma-aminobutyric acid
HDL: high-density lipoprotein
MPL: minimally processed lentil
MPP: minimally processed pork
RCT: randomized controlled trial
SPISE: single-point insulin sensitivity estimator.

